# Risk factors of chronic liver disease among adult patients in tertiary hospitals, Northern Ethiopia: An unmatched case-control study

**DOI:** 10.1101/2022.03.20.22272661

**Authors:** Migbar Sibhat, Tadesse Kebede, Dawit Aklilu

**Author notes:** Corresponding author: Migbar Sibhat, Affiliation: Dilla University, Dilla, Ethiopia.

## Abstract

**Introduction:** Chronic liver disease imposed substantial health and economic burden causing 1.75 million deaths and increased hospital admission rates. However, it is a neglected health issue in resource-limited countries like Ethiopia, with the paucity of data on the determinants of chronic liver disease. Besides, available studies considered single or limited factors. Thus, the aim of this study was to assess the determinant factors of chronic liver disease among patients who were attending at the gastroenterology units.

**Methods:** An unmatched case-control study was conducted on 281 (94 cases and 187 controls) systematically selected subjects at tertiary hospitals in Northern Ethiopia from November 2018 to June 2019. Data were collected using an interviewer-administered questionnaire and checklists, entered to Epi data version 4.4.2, and analyzed using SPSS version 25. Bi-variable and multivariate analyses were done using binary logistic regression. Variables with p< 0.25 in the bi-variable analysis were fitted to the final model. An adjusted odds ratio with 95% CI was used to declare significance and associations.

**Results:** An overall 281 (94 cases and 187 controls) patients attending at the gastroenterology units had been included with a response rate of 100%. Being chronic alcohol consumer (AOR=2.8; 95% CI: 1.1-7.0), having a history of herbal medication use (AOR=14; 95% CI: 5.2-42), history of injectable drug use (AOR=8.7; 95% CI: 3-24.8), and hepatitis B infection (AOR=12; 95% CI: 3.0-49) were found to have an independent association with chronic liver disease.

**Conclusion:** Alcohol consumption, history of herbal medication use, hepatitis B infection, and history of parenteral medication use were found to be determinant factors of chronic liver disease. Strengthening viral hepatitis screening activities, interventions focused on behavioral change, and linking traditional healers to the healthcare system is crucial.

**What is known about the subject?:** Chronic liver disease (CLD) causes substantial health and economic burden where more than 1 million deaths occurred due to CLD complications annually. Studies reported that CLD causes 1.8-10% of all deaths and responsible for increased hospital admission rates. From 1980 to 2010, chronic liver disease-related deaths doubled in sub-Saharan African countries. Despite measures have been designed and on implementation to decrease this burden, the access to these interventions is limited, and the number of patients increased dramatically.

**What does this study add?:**

- Different behavioral, socio-cultural, and clinical factors had a statistically significant association with chronic liver disease.
- Alcohol consumption, history of herbal medication use, hepatitis B infection (HBV^+^), and history of parenteral medication use were found to be determinant factors of chronic liver disease (CLD).

**Strength and limitations of the study:**

- Despite this study presented important findings that could input for the scientific world in the area of CLD, the study had its own limitations.
- Since almost all participants did not have documented medical checkups, it was challenging to extract previous history of chronic viral hepatitis. Hence, the effect of this pertinent variable was left unevaluated in this study.

## Introduction

Chronic liver disease (CLD) is defined as a progressive disease that causes deterioration and destruction of liver cells that can be diagnosed by using liver function test (LFT), imaging tests like Computed Tomography (CT scan), Magnetic Resonance Imaging (MRI), ultrasound, liver biopsy, and endoscopy(1-4). It can be classified as alcohol-induced, viral, autoimmune, metabolic, inherited, and fatty chronic liver diseases, of which alcoholic liver disease accounts for 48% of all deaths from chronic liver disease(5-7).

Globally, the liver disease accounts for more than 2 million deaths per year due to CLD, viral hepatitis, and Hepato-Cellular Carcinoma (HCC), where 1 million deaths were attributable to CLD complications(8-10). CLD causes substantial health and economic burden(11, 12). Studies reported that CLD causes 1.8-10% of all deaths and responsible for increased hospital admission rates, possibly due to an upwards shift in obesity and type 2 Diabetes Mellitus (DM)(13-16). From 1980 to 2010, chronic liver disease-related deaths doubled in sub-Saharan African countries(10).

Measures have been taken to decrease this burden by investing large amounts of money in funding research, vaccines and drug development, recruiting hepatologists and scientists. Moreover, the expanded program of immunization for HBV and mandatory HCV screening for blood transfusion were implemented to prevent new infections. However, access to these services is not achieved yet, and the number of patients increased dramatically (2, 17, 18).

Ethiopian reports indicated that about 12% of the hospital admissions and 31% of the mortality in medical wards were due to CLD(16). The Ethiopian Health and Nutrition Research Institute (EHNRI), Centers for Disease Control and Prevention (CDC), World Health Organization (WHO), and Tigray Regional Health Bureau established the CLD surveillance system in 2017(19). Despite these actions, CLD carried the highest burden, and remains a neglected health issue in low and middle-income countries(20). While studies were conducted in the developed world, data on determinant factors of CLD were scarce, especially in resource-limited countries, like Ethiopia. Furthermore, the available researches considered single or limited determinant factors of CLD where new variables such as herbal medication use were assessed in the current study. For instance, throughout our country, Ethiopia, just one study was conducted on this issue tried to investigate the effect of merely two factors (alcohol and viral hepatitis) on chronic liver diseases. Thus, the aim of this study was to assess the determinant factors of CLD among patients in the gastroenterology clinics.

## Methods and materials

### Study design, area and period

An institution-based unmatched case-control study was conducted among patients who had follow-up at gastroenterology units of tertiary hospitals in Northern Ethiopia from November 2018 to July 2019. Ayder comprehensive specialized hospital (ACSH), located in the Tigray region, is the second-largest tertiary hospital in Ethiopia next to Black Lion hospital, providing the gastroenterology services for more than 490 patients per month on average. The hospital had more than 80 specialist physicians, 29 sub-specialists, 740 nurses, numerous medical students and other health professionals serving more than 8 million people coming from Tigray, Afar, Southeastern parts of the Amhara region, and the Eritrean refugees.

### Study population

#### Cases

All patients confirmed as having CLD based on the diagnostic criteria by the duty physician and who were being treated in the gastroenterology clinics.

#### Controls

All patients declared as non-CLD (without CLD features) and being treated in the gastroenterology clinics.

### Sample size determination and sampling procedure

Epi info version 7 was used to determine the required sample size based on the following assumptions: 95% confidence level, 80% power, and 1:2 ratios of cases to controls. Male sex was considered to determine the sample size from the study conducted in Nigeria(21), which brought the largest sample size with percentage exposed among the controls=48.4%, percent of exposed among the cases=67.6%, and & adjusted odds ratio=2.2 that yields 255. After adding a 10% contingency, the total sample size required to conduct the study was 281 (94 cases and 187 controls). Then, a systematic random sampling technique was used to select both cases and controls. The sampling interval (k) was found by dividing the past two months of patient flow by the calculated sample size. An overall 980 patients (190 diagnosed as CLD and 790 non-CLD cases) were registered in the gastroenterology (GI) unit from September 1, 2018-October 30, 2018 that brought 190/94=2 for the cases and 790/187=4 for controls. Hence, after cascading by order of their GI clinic appointment schedule, every other CLD patient was included as a case, and every fourth non-CLD patient consecutive to the selected case was incorporated as a control. The first participant was selected randomly using the lottery method.

### Operational Definitions

#### Diagnostic Criteria

The diagnosis of CLD was declared using the combination of clinical features suggestive of CLD and either ultrasound findings consistent with CLD (presence of an irregular liver surface or liver parenchyma heterogeneity (22) or serological tests (The serum aspartate aminotransferase (AST) to platelet ratio index (APRI) using a threshold of 0.7 as an indicator of significant fibrosis. APRI was calculated by dividing the upper reference range of AST (U/L) to platelet count (10^9^/L) and multiplied by 100(23).

**Cases** (chronic liver disease) were patients who had confirmed CLD based on the diagnostic criteria by the duty physician and who were being treated in the gastroenterology clinics. **Controls** were those patients declared as non-CLD (without CLD features) and being treated in the gastroenterology clinics. **Alcohol consumption** was assessed by the CAGE standard screening tool. The tool has four yes/no questions where two and above yes responses were defined as alcoholic(22). **Body Mass Index (BMI)** was defined as the body mass (weight) divided by the square of the body height and was expressed in units of kg/m^**2**,^ and categorized to underweight (<18.5), normal (18.5–24.9), overweight (25.0–29.9), and obese (≥30.0)(23). **Thrombocytopenia (**low platelet levels in the blood) was classified as mild (75-150×10^3^ cells/µl), moderate (50-75×10^3^ cells/µl), and severe (<50×10^3^ cells/µl)(24).

### Data collection tools and procedures

Data were collected by face-to-face interview, chart review, and physical examination using a structured interviewer-administered questionnaire and checklists that were designed from different literature (21, 25, 26). The data collection tool comprised of socio-demographic variables, behavioral factors, socio-cultural factors, and clinical variables. Height (in meters) was taken using the hospital height scale while the patient is standing upright, and weight (in kilograms) was measured using standardized and calibrated weighing equipment. Three data collectors (BSc nurses) and one supervisor (MSc fellow) completed the data collection within eight weeks, from April 1 to May 30, 2019.

### Data quality assurance

The questionnaire was prepared in English, translated to local languages (Tigrigna and Amharic), and reverse translation was done to English to check the consistency. One week before the data collection, the questionnaire was pre-tested on 5% of the samples (15 patients) that were not incorporated in the actual data for analysis by the principal investigator to assure the consistency and clarity of the tool where unclear items were modified accordingly. One day training was given for data collectors and the supervisor about the research objective, eligible study subjects, data collection tools and procedures, and interview methods. Data collection was coordinated and appraised by the supervisor and principal investigator.

### Data processing and analysis

Data were cleaned and entered using Epi data version 4.4.2 and analyzed using SPSS version 25. An exploratory analysis was carried out to determine the nature of data, such as normality and the presence of outliers as well as the level of missing values. Then, the data were described using relative frequency, percent, mean with standard deviation. Binary logistic regression was used to conduct both bi-variable and multivariate analysis. The model fitness to the data was checked using Hosmer and Lemeshow test, and multi-collinearity was investigated using the variance inflation factor. In the bi-variable analysis, variables with p-values<0.25 were scrutinized and fitted to multivariate analysis to identify the independent effects of each covariate on the outcome variable. In the final logistic regression model, statistical significance was declared at p*<*0.05, and the presence and strength of associations were summarized using an adjusted odds ratio with 95% confidence intervals. Finally, study findings were displayed in texts and tables.

### Patient and public involvement

Patients or the public did not take part in the study design, conduct, reporting, or dissemination plans of this research.

### Ethical Considerations

The study was conducted after approved by Institutional Review Board (IRB) of Mekelle University, Health Sciences College (exemption ID=ERC 1258/2019). A letter of cooperation was received from the School of Nursing, and permission to conduct the study was offered from the chief clinical director, matron officer, and gastroenterology unit coordinators. The study was conducted per the declaration of Helsinki. Before data collection, the purpose and objective of the study were described to the study participants, and verbal and written informed consent was obtained. Besides, respondents were informed to terminate the interview any time. Confidentiality of the information was secured throughout the study process using code instead of any personal identifier & is meant only for the study.

## Results

An overall 281 (94 cases and 187 controls) patients who had follow-up at gastroenterology units participated in this study with a response rate of 100%.

### Socio-demographic characteristics

The current study finding showed that 77 (81.9%) cases and 115 (61.5%) of controls were males. The mean (SD) age of the study subjects who had chronic liver diseases was 41±13 years for cases with a maximum of 18 to 75 years, whereas 40±15 years among controls. The age distribution showed that 35 (37.2%) of cases and 56 (29.9%) controls were between the age group of 30 to 41 years, in which 38.5% of participants in this age group had CLD. One-hundred eleven (39.5%) participants came from rural areas, of that 37.8% were diagnosed with CLD. The proportion of CLD among participants who were soldiers, farmers, and drivers was 44.4%, 41.1%, and 30.8% sequentially. Again, of the 23 (8.2%) participants who were unemployed, eight (34.8%) had diagnosed with CLD. Of the participants, 67 (71.3%) cases and 113 (60.4%) controls were married. On the other hand, 28.7% of the study subjects having chronic liver diseases could not read and write (Table 1).

**Table 1:** Distribution of socio-demographic characteristics among patients who had been attending at the gastroenterology units of tertiary hospitals, Northern Ethiopia, 2019 (N=281)

| Independent variables | Category | Diseases status |  | Total (%) |
| --- | --- | --- | --- | --- |
|  |  | Cases (%) | Controls (%) |  |
| **Sex** | Male | 77 (81.9) | 115 (61.5) | 192 (68.3) |
|  | Female | 17 (18.1) | 72 (38.5) | 89 (31.7) |
| **Age(years)** | 18-29 | 18 (19.1) | 52 (27.8) | 70 (24.9) |
|  | 30-41 | 35 (37.2) | 56 (29.9) | 91 (32.4) |
|  | 42-53 | 22 (23.4) | 43 (23.0) | 65 (23.1) |
|  | 54-65 | 15 (16.0) | 25 (13.4) | 40 (14.2) |
|  | >65 | 4 (4.3) | 11 (5.9) | 15 (5.3) |
| **Residence** | Urban | 52 (55.3) | 118 (63.1) | 170 (60.5) |
|  | Rural | 42 (44.7) | 69 (36.9) | 111 (39.5) |
| **Occupation** | Farmer | 37 (39.4) | 53 (28.3) | 90 (32.0) |
|  | Driver | 4 (4.3) | 9 (4.8) | 13 (4.6) |
|  | Military | 4 (4.3) | 5 (2.7) | 9 (3.2) |
|  | Housewife | 9 (9.6) | 34 (18.2) | 43 (15.3) |
|  | Unemployed | 8 (8.5) | 15 (8.0) | 23 (8.2) |
|  | Government employee | 28 (29.8) | 60 (32.1) | 88 (31.3) |
|  | Student | 4 (4.3) | 11 (5.9) | 15 (5.3) |
| **Marital status** | Single | 20 (21.3) | 60 (32.1) | 80 (28.5) |
|  | Married | 67 (71.3) | 113 (60.4) | 180 (64.1) |
|  | Widowed | 4 (4.3) | 5 (2.7) | 9 (3.2) |
|  | Divorced | 3 (3.2) | 9 (4.8) | 12 (4.3) |
|  | Can't read and write | 27 (28.7) | 52 (27.8) | 79 (28.1) |
| **Educational status** | Read and write | 13 (13.8) | 21 (11.2) | 34 (12.1) |
|  | Primary education | 26 (27.7) | 33 (17.6) | 59 (21.0) |
|  | Secondary education | 15 (16.0) | 34 (18.2) | 49 (17.4) |
|  | College and above | 13 (13.8) | 47 (25.1) | 60 (21.4) |
| **Average monthly** | <2000Birr | 66 (70.2) | 88 (47.1) | 154 (54.8) |
| **income** | $\geq 2000$birr | 28 (29.8) | 99 (52.9) | 127 (45.2) |

### The Behavioral and Socio-cultural Characteristics of the Study Participants

The study finding revealed that the proportion of drinkers (CAGE≥2) among cases (CLD) 53 (51%) was higher than among controls 51 (49%). Also, 33 (62.3%) cases and 21 (41.2%) controls were consuming alcohol for≥10years. The BMI of study participants ranged from 17.5-29.1kg/m^2^ with a mean (SD) value of 22±3 kg/m^2^ among cases where it was 21.7±3 kg/m^2^ among controls with the minimum of 17.5 kg/^m2^ and maximum 28.2 kg/m^2^. Regarding the body tattoo, the proportion of patients having body tattoo was higher among CLD patients 34 (58.6%) than controls 24 (41.4). The study finding also reported that 119 (42.3%) participants used herbal medications, of which 77 (81.9%) were among cases and 42 (22.5%) among controls, whereas 29 (31%) cases and 42 (22.5%) controls were overweight (25-29.9 kg/m^2^) (Table 2).

**Table 2:** Behavioral and socio-cultural characteristics of patients who had been attending at the gastroenterology units of tertiary hospitals, Northern Ethiopia, 2019 (N=281)

| Independent variables | Category | Diseases status |  | Total (%) |
| --- | --- | --- | --- | --- |
|  |  | Cases (%) | Control (%) |  |
| **Alcohol drinking (CAGE Score)** | Yes ($\geq 2$) | 53 (56.4) | 51 (27.3) | 104 (37.0) |
|  | No (<2) | 41 (43.6) | 136 (72.7) | 177 (63.0) |
| **Duration of alcohol drinking** | <10 years | 20 (37.7) | 30 (58.8) | 50 (48.0) |
| | $\geq 10$ years | 33 (62.3) | 21 (41.2) | 54 (51.9) |
| **Cigarette smoking** | Yes | 2 (2.1) | 3 (1.6) | 5 (1.8) |
|  | No | 92 (97.9) | 184 (98.4) | 276 (98.2) |
| **BMI(Kg/m<sup>2</sup>)** | <18.5 | 16 (17.0) | 47 (25.1) | 63 (22.4) |
|  | 18.5-24.9 | 48 (51.1) | 94 (50.3) | 142 (50.5) |
|  | 25-29.9 | 29 (30.9) | 42 (22.5) | 71 (25.3) |
|  | >30 | 1 (1.1) | 4 (2.1) | 5 (1.8) |
| **Tattoo on the body** | Yes | 34 (36.2) | 24 (12.8) | 58 (20.6) |
|  | No | 60 (63.8) | 163 (87.2) | 223 (79.4) |
| **Herbal medication use** | Yes | 77 (81.9) | 42 (22.5) | 119 (42.3) |
|  | No | 17 (18.1) | 145 (77.5) | 162 (57.7) |

**Abbreviations:** CAGE, cut down, annoyed by criticism, guilty about drinking, eye-opener drinks (a test for alcoholism); BMI, body mass index; Kg/m<sup>2</sup>, kilograms per meter square

### The Clinical Characteristics of the Study Participants

Almost half (48%) of the respondents had a history of parenteral medication use where the proportion of use was higher 78 (58.2%) among patients diagnosed with chronic liver diseases than 56 (41.8%) among those not having chronic liver diseases. The mean (SD) of AST level was 66±74IU/L and ranging from 12.6 to 368.0IU/L for cases and 48± 58 IU/Land ranging from 9 to 462IU/L for controls. Thirty-nine (14%) of the patients had AST level of ≥80, of which 23 (59%) were the cases, and 16 (41%) were the controls. Whereas 26 (9.2) study subjects had severe thrombocytopenia, of which 23 (88.5%) were cases, and three (11.5%) were the controls. Sixty (63.8%) cases and 30 (16%) controls had a personal history of liver diseases. Moreover, 35 (12.5%) patients had a history of surgery, and 32 (11.4%) tested positive for serum HCV. About 36 (12.8%), 41(14.6%), and 52(18.5%) had type II diabetes mellitus, a history of blood transfusion, and a family history of liver diseases consecutively (Table 3).

**Table 3:**
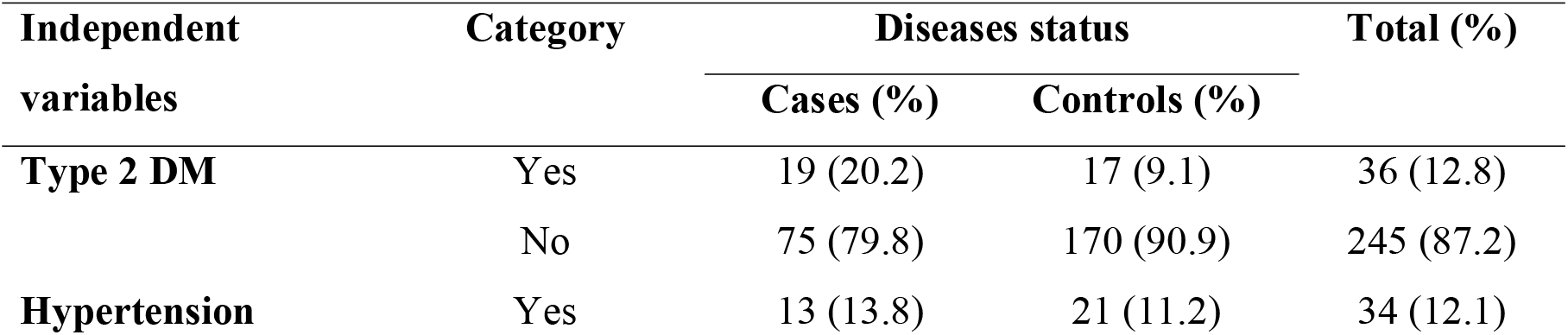
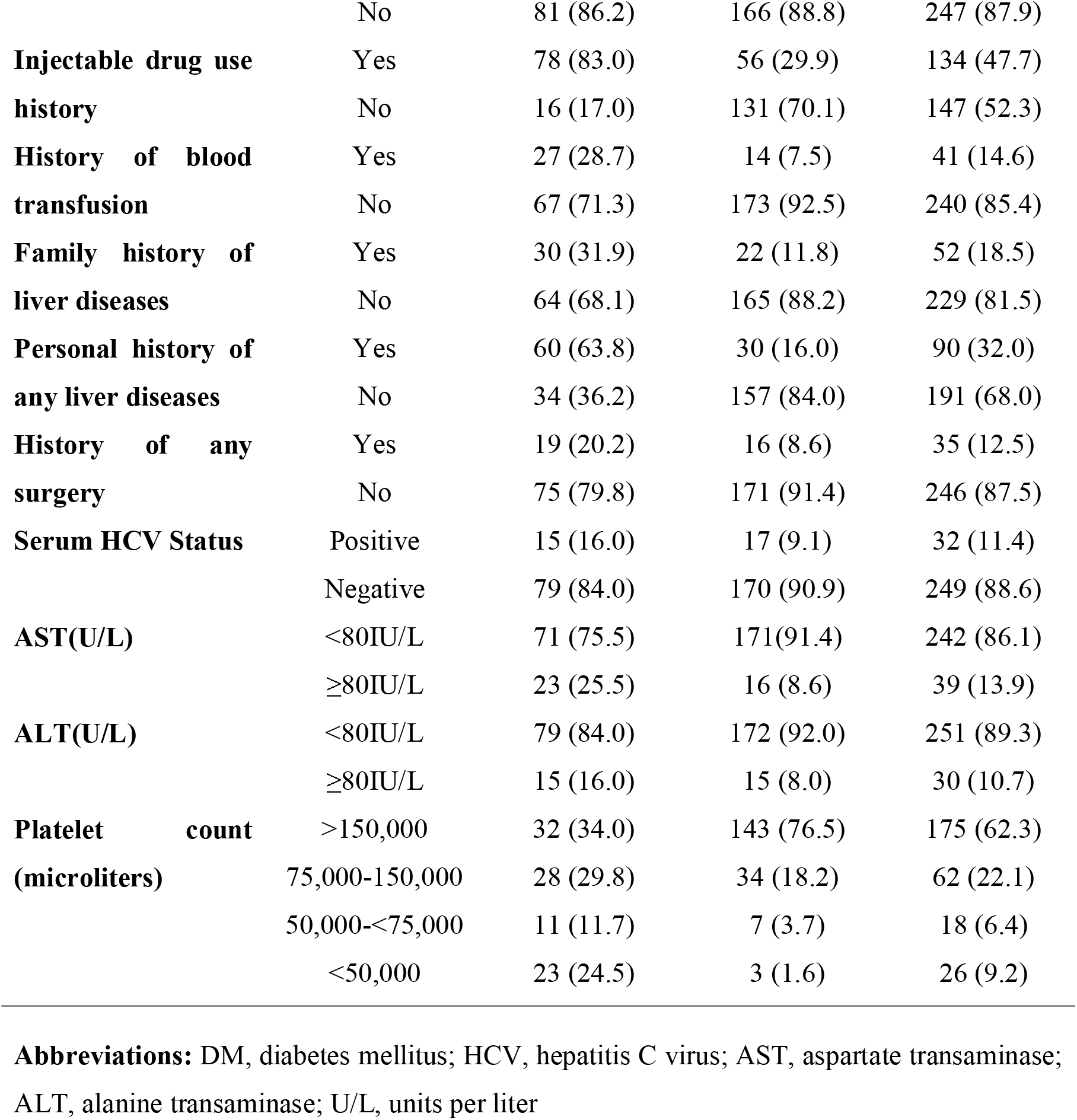
Distribution of clinical and laboratory-related characteristics among patients who had been attending at the gastroenterology units of tertiary hospitals, Northern Ethiopia, 2019 (N=281)

| Independent variables | Category | Diseases status |  | Total (%) |
| --- | --- | --- | --- | --- |
|  |  | Cases (%) | Controls (%) |  |
| **Type 2 DM** | Yes | 19 (20.2) | 17 (9.1) | 36 (12.8) |
|  | No | 75 (79.8) | 170 (90.9) | 245 (87.2) |
| **Hypertension** | Yes | 13 (13.8) | 21 (11.2) | 34 (12.1) |
|  | No | 81 (86.2) | 166 (88.8) | 247 (87.9) |
| **Injectable drug use history** | Yes | 78 (83.0) | 56 (29.9) | 134 (47.7) |
|  | No | 16 (17.0) | 131 (70.1) | 147 (52.3) |
| **History of blood transfusion** | Yes | 27 (28.7) | 14 (7.5) | 41 (14.6) |
|  | No | 67 (71.3) | 173 (92.5) | 240 (85.4) |
| **Family history of liver diseases** | Yes | 30 (31.9) | 22 (11.8) | 52 (18.5) |
|  | No | 64 (68.1) | 165 (88.2) | 229 (81.5) |
| **Personal history of any liver diseases** | Yes | 60 (63.8) | 30 (16.0) | 90 (32.0) |
|  | No | 34 (36.2) | 157 (84.0) | 191 (68.0) |
| **History of any surgery** | Yes | 19 (20.2) | 16 (8.6) | 35 (12.5) |
|  | No | 75 (79.8) | 171 (91.4) | 246 (87.5) |
| **Serum HCV Status** | Positive | 15 (16.0) | 17 (9.1) | 32 (11.4) |
|  | Negative | 79 (84.0) | 170 (90.9) | 249 (88.6) |
| **AST(U/L)** | <80IU/L | 71 (75.5) | 171(91.4) | 242 (86.1) |
|  | ≥80IU/L | 23 (25.5) | 16 (8.6) | 39 (13.9) |
| **ALT(U/L)** | <80IU/L | 79 (84.0) | 172 (92.0) | 251 (89.3) |
|  | ≥80IU/L | 15 (16.0) | 15 (8.0) | 30 (10.7) |
| **Platelet count (microliters)** | >150,000 | 32 (34.0) | 143 (76.5) | 175 (62.3) |
|  | 75,000-150,000 | 28 (29.8) | 34 (18.2) | 62 (22.1) |
|  | 50,000-<75,000 | 11 (11.7) | 7 (3.7) | 18 (6.4) |
|  | <50,000 | 23 (24.5) | 3 (1.6) | 26 (9.2) |

**Abbreviations:** DM, diabetes mellitus; HCV, hepatitis C virus; AST, aspartate transaminase; ALT, alanine transaminase; U/L, units per liter

### Risk factors of chronic liver diseases

After data description, the bi-variable analysis was conducted using the binary logistic regression model to determine the variables that should be fitted to the final model for multivariate analysis. During the bi-variable analysis, being male, rural residence, educational status, alcohol drinkers, tattoo on the body, history of herbal medication use, history of Type2 DM, history of parental medication use, family history of liver diseases, personal-history of liver diseases, positive serum HBV and HCV were statistically significant at P-value <0.25 and were entered to the final logistic regression model.

Finally, in the multivariate analysis, being alcohol consumer (CAGE score ≥2), history of herbal medication use, history of parenteral medication use, and being HBV-positive showed statistically significant association with CLD at p-value ≤0.05 (Table 4).

**Table 4:**
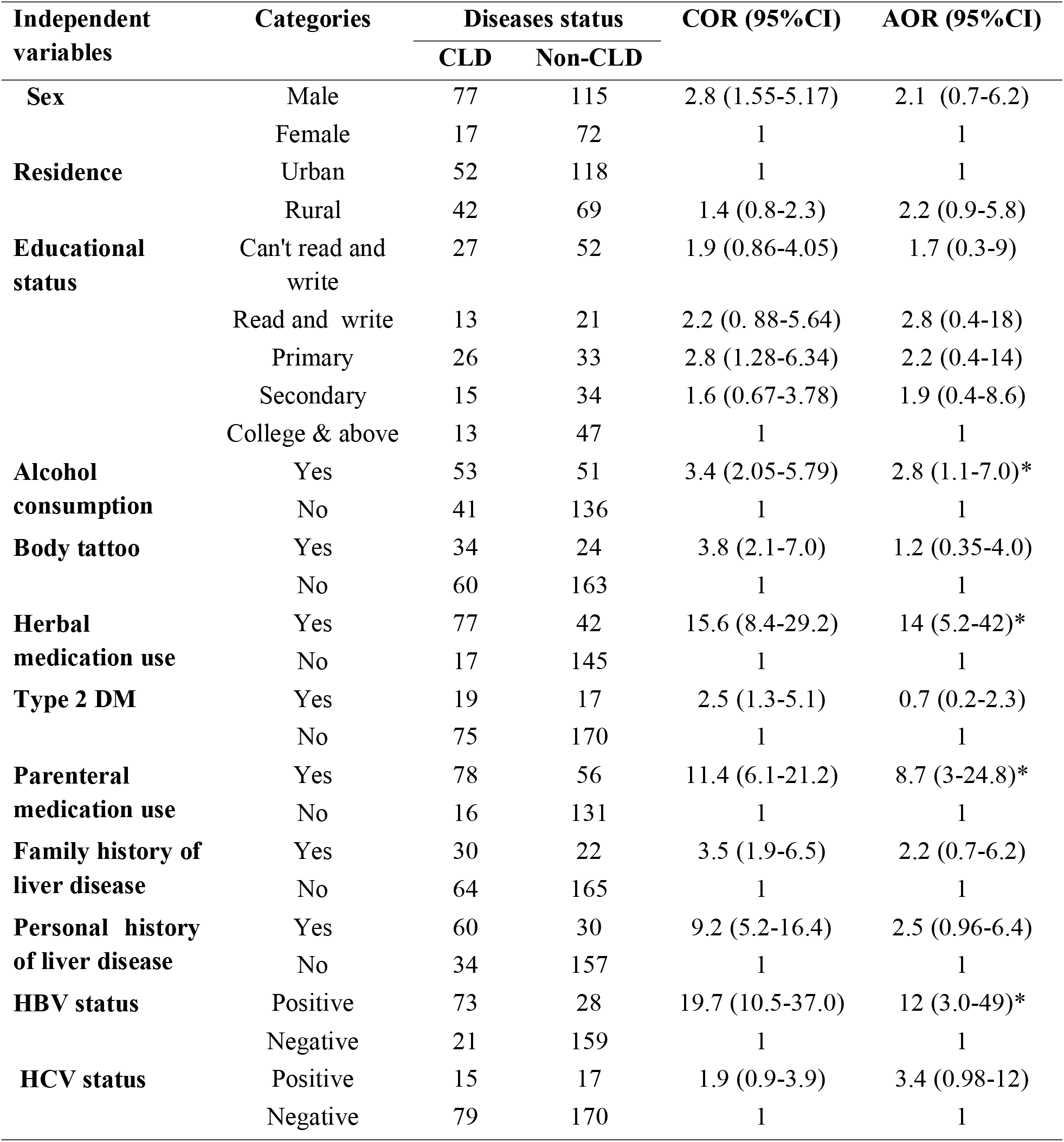
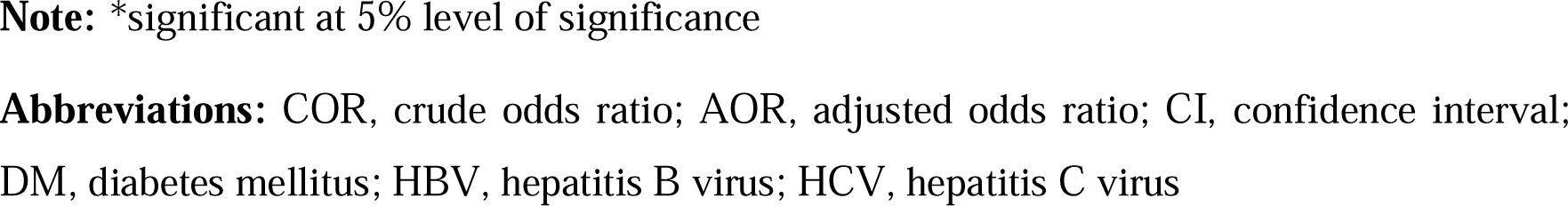
Bi-variable and multivariate logistic regression analysis for determinants of CLD among patients who had been attending at the gastroenterology units of tertiary hospitals, Northern Ethiopia, 2019 (N=281)

| Independent variables | Categories | Diseases status |  | COR (95%CI) | AOR (95%CI) |
| --- | --- | --- | --- | --- | --- |
|  |  | CLD | Non-CLD |  |  |
| **Sex** | Male | 77 | 115 | 2.8 (1.55-5.17) | 2.1 (0.7-6.2) |
|  | Female | 17 | 72 | 1 | 1 |
| **Residence** | Urban | 52 | 118 | 1 | 1 |
|  | Rural | 42 | 69 | 1.4 (0.8-2.3) | 2.2 (0.9-5.8) |
| **Educational status** | Can't read and write | 27 | 52 | 1.9 (0.86-4.05) | 1.7 (0.3-9) |
|  | Read and write | 13 | 21 | 2.2 (0.88-5.64) | 2.8 (0.4-18) |
|  | Primary | 26 | 33 | 2.8 (1.28-6.34) | 2.2 (0.4-14) |
|  | Secondary | 15 | 34 | 1.6 (0.67-3.78) | 1.9 (0.4-8.6) |
|  | College & above | 13 | 47 | 1 | 1 |
| **Alcohol consumption** | Yes | 53 | 51 | 3.4 (2.05-5.79) | 2.8 (1.1-7.0)* |
|  | No | 41 | 136 | 1 | 1 |
| **Body tattoo** | Yes | 34 | 24 | 3.8 (2.1-7.0) | 1.2 (0.35-4.0) |
|  | No | 60 | 163 | 1 | 1 |
| **Herbal medication use** | Yes | 77 | 42 | 15.6 (8.4-29.2) | 14 (5.2-42)* |
|  | No | 17 | 145 | 1 | 1 |
| **Type 2 DM** | Yes | 19 | 17 | 2.5 (1.3-5.1) | 0.7 (0.2-2.3) |
|  | No | 75 | 170 | 1 | 1 |
| **Parenteral medication use** | Yes | 78 | 56 | 11.4 (6.1-21.2) | 8.7 (3-24.8)* |
|  | No | 16 | 131 | 1 | 1 |
| **Family history of liver disease** | Yes | 30 | 22 | 3.5 (1.9-6.5) | 2.2 (0.7-6.2) |
|  | No | 64 | 165 | 1 | 1 |
| **Personal history of liver disease** | Yes | 60 | 30 | 9.2 (5.2-16.4) | 2.5 (0.96-6.4) |
|  | No | 34 | 157 | 1 | 1 |
| **HBV status** | Positive | 73 | 28 | 19.7 (10.5-37.0) | 12 (3.0-49)* |
|  | Negative | 21 | 159 | 1 | 1 |
| **HCV status** | Positive | 15 | 17 | 1.9 (0.9-3.9) | 3.4 (0.98-12) |
|  | Negative | 79 | 170 | 1 | 1 |

**Note:** \*significant at 5% level of significance
**Abbreviations:** COR, crude odds ratio; AOR, adjusted odds ratio; CI, confidence interval; DM, diabetes mellitus; HBV, hepatitis B virus; HCV, hepatitis C virus

The study finding showed that alcohol drinkers (CAGE score ≥2) were 2.8 times (AOR=2.8; 95% CI: 1.1-7.0) more likely to develop CLD compared to non-drinkers. The present study also revealed that the risk of developing CLD among clients who had a history of herbal medication use was higher (AOR=14; 95% CI: 5.2-42) than patients who had no history of herbal medication use. Furthermore, the odds of patients having a history of parenteral medication use (injectable drug use (IDU) to experience chronic liver disease (CLD) was 8.7 times (AOR=8.7; 95% CI: 3-24.8) greater than their counterparts. The study result also notified that patients who had tested positive for hepatitis B virus (HBV^+^) were 12 times (AOR=12; 95% CI: 3.0-49) at an increased risk of developing CLD compared to those who had negative serum HBV status.

## Discussion

The current study intended to investigate the factors that determine chronic liver diseases. According to the study result, being an alcohol drinker, being HBV-positive, having a history of herbal medication use, and having a history of parenteral medication use were independent predictors of chronic liver diseases.

The study finding showed that alcohol drinkers (CAGE score ≥2) were 2.8 times (AOR=2.8; 95% CI: 1.1-7.0) more likely to develop CLD compared to non-drinkers, which was in line with the study conducted in Iran, Taiwan, USA, Europe, Italy, and Addis Ababa(21, 22, 26-28). The possible explanation could be liver processes over ninety percent of consumed alcohol. Chronic alcohol drinking predisposes to alcoholic hepatitis and destruction of liver cells, which results in cellular mutation of the liver and scarring of the liver cells (cirrhosis). That, in turn, could lead to chronic liver disease and hepatic cell carcinoma (HCC)(29, 30).

Nevertheless, the result of this study was inconsistent with the study conducted in Nigeria(25). The discrepancy might be due to the criteria used to classify drinker and non-drinker. We and other previous studies (24) used the CAGE standard tool, which cannot consider the amount of alcohol ingested. On the contrary, the Nigerian study used years of drinking and amount of alcohol consumed to classify drinkers vs. non-drinker. As it has been shown that the amount of alcohol ingested (irrespective of the type of alcohol consumed) was the most important determinant factors for CLD, and the risk of developing the diseases increased with the ingestion of >60-80grams per day for ten years in males and >20g/day in females. Therefore, the variation might be due to this reason.

The present study also revealed that the risk of developing CLD among clients who had a history of herbal medication use was higher (AOR=14; 95% CI: 5.2-42) than patients who had no history of herbal medication use. Earlier studies also reported that history of herbal medication use increased risk of CLD(25, 31). This could be explained that herbal products are not tested with the scientific approval required for conventional drugs and cannot be marketed for the diagnosis, treatment, cure or prevention of the diseases. One of the roles of the liver is to act as a filter for toxins through a complex metabolic process by taking the non-toxic components and flushing toxins out of the body. Certain herbs can damage the liver cells during this process since they can form toxic metabolites(32).

Furthermore, the odds of patients having a history of parenteral medication use (injectable drug use (IDU) to experience chronic liver disease (CLD) was 8.7 times (AOR=8.7; 95% CI: 3-24.8) greater than their counterparts. This finding was supported by previous studies conducted in Nigeria and Iran(25, 26). The most likely rationale for this could be due to the effect of injectable drugs on the liver. Drug-induced chronic liver diseases may result from the direct toxicity of metabolites of administered drugs or due to immune-mediated mechanisms. That can be enhanced further by the subsequent inflammatory reaction. Drug metabolism may generate bioactive products that interact with various cellular organelles, such as mitochondria, leading to hepatocyte dysfunction and cellular demise(33).

On the other hand, the findings of the study conducted in Iran(34) contradict with our study result. This difference might be due to the frequent use of parenteral medication in developing countries like Ethiopia for empirical therapy due to the lack of confirmatory diagnostic options such as culture. Besides, many patients self-medicate since most drugs are available without the physician’s prescription. The other possible reason might be the difference in the sample size used.

The study result also notified that patients who had tested positive for hepatitis B virus (HBV^+^) were 12 times (AOR=12; 95% CI: 3.0-49) at an increased risk of developing CLD compared to those who had negative serum HBV status. This report agreed with the findings of former studies done in Europe, Asia, and Africa (21, 37, 38). The scientific justification might be that histological ends of chronic active HBV could lead to liver cell necrosis, inflammation, cytokine production, and liver scarring (fibrosis). In other words, T-lymphocytes-mediated cytotoxicity that probably directed against Hepatitis B antigen on the hepatocellular membrane has a leading role in hepatocyte cytolysis. It may also be linked with HBV-DNA integration into hepatocyte clones and autoimmune-like reactions leading to further liver cell damage(29). Despite this study presented important findings that could input for the scientific world in the area of CLD, the study had its own limitations. Since almost all participants did not have documented medical checkups, it was challenging to extract previous history of chronic viral hepatitis. Hence, the effect of this pertinent variable was left unevaluated in this study.

## Conclusion

This study notified that different behavioral, socio-cultural, and clinical factors had a statistically significant association with chronic liver diseases. Alcohol consumption, history of herbal medication use, hepatitis B infection (HBV^+^), and history of parenteral medication use were found to be determinant factors of chronic liver disease (CLD).

## Data Availability

Extra data that support the findings of this study are available and can be shared upon reasonable and legal request via

## Abbreviations

AORs: Adjusted odds ratios
CIs: confidence intervals
CLD: Chronic liver disease
HCC: Hepato-Cellular Carcinoma
ACSH: Ayder comprehensive specialized hospital
DM: diabetes mellitus
CAGE: cut down, annoyed by criticism, guilty about drinking, eye-opener drinks (a test for alcoholism)
BMI: body mass index
HBV: hepatitis B virus
HCV: hepatitis C virus
WHO: World Health Organization

## Acknowledgments

Study participants, data collectors, supervisors, hospital staff, and administrators were acknowledged for their unfailing contribution. The authors would also like to thank Mekelle University that covers the financial requirements for data collectors.

## Authors’ contributions

TK, MS & DA participated in the study conception and proposal development as well as collection and interpretation of data. MS, TK, DA & participated in data entry, cleaning, and performed the statistical analysis. MS drafted, compiled, edited and formatted the manuscript for publication. All authors read, critically revised, and approved the final manuscript and agreed to be accountable for all aspects of the work.

## Consent for publication

Not applicable

